# Temporal stability and state-dependence of retrospective self-reports of childhood maltreatment in major depression: a two-year longitudinal analysis of the childhood trauma questionnaire

**DOI:** 10.1101/2021.08.31.21262884

**Authors:** Janik Goltermann, Susanne Meinert, Carina Hülsmann, Katharina Dohm, Dominik Grotegerd, Ronny Redlich, Lena Waltemate, Hannah Lemke, Katharina Thiel, David M. A. Mehler, Verena Enneking, Tiana Borgers, Jonathan Repple, Marius Gruber, Nils Winter, Tim Hahn, Katharina Brosch, Tina Meller, Kai G. Ringwald, Simon Schmitt, Frederike Stein, Julia-Katharina Pfarr, Axel Krug, Igor Nenadić, Tilo Kircher, Nils Opel, Udo Dannlowski

## Abstract

**Background:** Retrospective self-reports of childhood maltreatment are widely used in research and clinical practice. However, their validity has been questioned due to potential depressive bias. Yet, systematic investigations of this matter are sparse. Thus, we investigate if and to what extent retrospective maltreatment reports vary in relation to longitudinal changes in depressive symptomatology.

**Methods:** Two-year temporal stability of maltreatment reports was assessed via the Childhood Trauma Questionnaire (CTQ). Diagnosis and depressive symptoms were assessed using clinical interview and the Beck Depression Inventory. From two independent cohorts (MACS and MNC) we included a total of n=347 major depressive disorder (MDD) patients, n=419 healthy controls (HC), and a subsample with a lifetime first depressive episode between both assessments (n=27). Analysis plan and hypotheses were preregistered prior to data sighting.

**Results:** Maltreatment reports were highly stable in HC and MDD patients across both cohorts (ICC=.956; 95%-CI[.949, .963] and ICC=.950; 95%-CI[.933, .963]) and temporal stability did not differ between groups. Stability was lower for cutoff-based maltreatment categorization (K=.551; 95%-CI[.479, .622] and K=.507; 95%-CI[.371, .640]). Baseline maltreatment reports were associated with concurrent and future depression severity. However, longitudinal changes in depression severity predicted variability in CTQ scores only to a small extent across cohorts (*b*=0.101, *p*=.009, R^2^=.021 and *b*=0.292, *p*=.320), with the effect being driven by emotional maltreatment subscales.

**Conclusions:** Findings suggest that the CTQ provides temporally stable self-reports of childhood maltreatment in healthy and depressed populations, and are only marginally biased by depressive symptomatology. Gradual rather than categorical conceptualization of maltreatment is advised for improving psychometric quality.

## Introduction

Childhood maltreatment (CM) has been implicated as a major risk factor for incidence and trajectory of depression and other psychiatric disorders (Lippard & Nemeroff, 2020). As such, maltreatment is often referred to as one of the key variables necessary to utilize for personalized medicine in psychiatry (Prendes-Alvarez & Nemeroff, 2016). To that end, a valid and practically feasible assessment of CM is crucial. However, the validity of currently established assessment methods has frequently been questioned (Hardt, Sidor, Bracko, & Egle, 2006).

Due to its extensive relevance, the consequences of CM have been subject of a plethora of research. However, the majority of empirical evidence relies on observational designs that implicate inherent limitations concerning their causal conclusiveness. Increased risk for the development of psychiatric disorders is among the most cited effects of CM, while most studies supporting this evidence rely on retrospective self-report measures (Green et al., 2010; Putnam, Harris, & Putnam, 2013) due to their high practicability (Hovdestad, Campeau, Potter, & Tonmyr, 2015). Cross-sectional studies cannot preclude that effects are at least partly inflated by psychopathological mechanisms affecting the recall and/or report of CM. This could particularly be true for Major Depressive Disorders (MDD). A negativity bias in regard to the appraisal of oneself, one’s future, and one’s environment is a core feature in MDD and central to influential cognitive models of depression (Beck, Rush, Shaw, & Emery, 1987). Correspondingly, negative biases have been described in different aspects of memory processes such as retrospective recall of mood state and autobiographical memories in depressed participants (Sato & Kawahara, 2011) and in healthy participants with experimentally induced low mood (Au Yeung, Dalgleish, Golden, & Schartau, 2006). It has been suggested that retrospective reports of CM could be biased in a similar way by depressive symptoms (Hardt & Rutter, 2004).

The suspected bias of CM reports due to depressive symptoms and the common use of retrospective self-reports in research make it crucial to re-evaluate our knowledge in this field. However, systematic assessments of the susceptibility of retrospective CM measures to depressive states are sparse. Fergusson and colleagues investigated temporal stability of retrospective interview regarding abuse at the ages of 18 and 21 in a population sample. They found that CM categorization was fairly unstable over the three-year interval but this variance was not systematically associated with changes in psychiatric status (Fergusson, Horwood, & Woodward, 2000). A major limitation of this study is, that it is unclear how many participants with clinically relevant symptoms of depression were included. Further it is questionable if findings generalize to CM assessments using well-established self-report instruments, and to other age groups. The latter is relevant because retrospective measures of CM are not only used in adolescents and young adults but throughout a wide age range up to participants over 60 years of age (Green et al., 2010).

One of the most widely used retrospective self-report questionnaires assessing CM is the Childhood Trauma Questionnaire (CTQ) (Bernstein et al., 1994), which has been utilized in hundreds of studies investigating CM (Viola et al., 2015). The psychometric qualities of the CTQ have been demonstrated in various patient samples (e.g. Hernandez et al., 2013; Spinhoven et al., 2014). and occasional evidence of high test-retest reliability exists (Cammack et al., 2017). However, although the latent construct measured by the CTQ naturally should be stable over time, the test-retest reliability seems to decreases with increasing duration of test-retest intervals (Bernstein et al., 1994; Cammack et al., 2017). Thus, it appears evident that significant variation in self-reports of CM over time is present. However, it is unknown to what extent this temporal variation can be explained by current depressive symptoms.

Validating any measure of CM is difficult due to a lack of a flawless measure it could be validated with. Prospective and often more objective measures (e.g. child protective services or hospital- or court records) have shown little overlap with retrospective self-report data (Baldwin, Reuben, Newbury, & Danese, 2019). However, that may be due to a limited sensitivity of prospective measures particularly regarding emotional maltreatment that is difficult to objectify or is not officially reported for other reasons (Hardt & Rutter, 2004).

CM has frequently been discussed as a key variable for the utilization in personalized medicine as lower treatment outcomes in maltreated individuals call for customized interventions for this patient group (Prendes-Alvarez & Nemeroff, 2016). However, in the context of personalized medicine, retrospective reports of CM are arguably without an alternative method of assessment as prospective measures are of virtually no use for the practical application of tailored treatments. It would thus be highly relevant for clinicians and researchers to know if and to what extent retrospective assessment of CM could be biased by concurrent depressive symptoms.

Hence, our main goal is to assess the extent of a depressive bias on retrospective self-reports of CM using the CTQ, and the validation of an approach adjusting CTQ scores for depressive symptoms. Therefore, existing data derived from a longitudinal design is used to investigate associations of changes in retrospective CM reports with changes in depression severity.

First, we aim to assess the test-retest reliability of retrospective self-reports of CM over a two-year interval. Further, we test the hypotheses that: 1) CM self-reports are biased by acute depressive symptoms and that 2) its reliability can be improved by adjusting CM reports for depressive symptom severity.

In addition, we investigate to what extent the susceptibility of longitudinal CTQ assessments to depressive symptom changes is differentially influenced across different maltreatment subtypes, depending on the age at assessment, across absolute levels of depression severity, or by the categorization of CM experiences (maltreated vs. non-maltreated).

## Methods

### Participants and design

Data sets from two independent cohorts were combined: the Marburg-Münster-Affective-Disorders-Cohort-Study (MACS) (Kircher et al., 2019) and the Münster-Neuroimaging-Cohort (MNC) (Opel et al., 2019). Both studies are ongoing and comprise healthy control (HC) and MDD participants, with age 18-65 years. A longitudinal design was applied with a baseline assessment (T_1_) and a follow-up assessment (T_2_) of all measures two to three years later (test-retest interval: MACS: M=776.5 days[SD=86.3]; MNC: M=816.4 days[SD=107.8]).

In the MACS, patients with acute or remitted MDD episodes were included irrespective of current treatment. In the MNC cohort all MDD patients underwent inpatient treatment for a current depressive episode at study inclusion. For both cohorts participants that have never had a psychiatric disorder at T_1_ and fulfill the diagnostic criteria for a depression within the 2-year interval were excluded from our main analyses and were investigated in a pooled converter sample (pooling converters from both cohorts for statistical power). A flowchart and detailed description of all data exclusion steps for both samples is provided in the supplements (Fig S1) and the supplementary methods section. The final samples included n=271 MDD and n=327 HC within MACS, and n=76 MDD and n=92 HC within MNC. Participants in MACS/MNC were on average 35.41/40.98 years old (SD 13.38/11.42) and comprised 389/84 females.

### Ethical Considerations

The MNC was approved by the ethics committee of the Medical Faculty of University of Münster (2007-307-f-S). The MACS was approved by the Ethics Committees of the Medical Faculties, University of Marburg (AZ: 07/14) and University of Münster (AZ: 2014-422-b-S). The study was performed in accordance with the ethical guidelines and regulations. All participants gave written informed consent prior to examination and received financial compensation.

### Procedure and measures

The German 28-item version of the Childhood-Trauma-Questionnaire (CTQ) (Wingenfeld et al., 2010) was administered to assess CM experienced before age 18. The CTQ is a retrospective self-report questionnaire assessing emotional neglect, physical neglect, emotional abuse, physical abuse and sexual abuse, as well as a denial subscale (Bernstein et al., 1994). The sum of subscale scores can be used to represent an overall maltreatment load. Categorical cutoffs were used to dichotomize participants into maltreated and non-maltreated for additional analyses (Walker et al., 1999).

Current symptomatology was assessed using the self-report questionnaire Beck-Depression-Inventory (BDI) (Beck, Ward, Mendelson, Mock, & Erbaugh, 1961). Changes in continuous CTQ and BDI scores from T_1_ to T_2_ are calculated as CTQ_T1_-CTQ_T2_=ΔCTQ and BDI_T1_-BDI_T2_=ΔBDI. As an alternate measure for depression severity, the 21-item Hamilton-Depression-Rating-Scale (HDRS) (Hamilton, 1960) was applied by trained raters.

Psychiatric diagnoses or the lack thereof was confirmed by trained personnel using the Structural-Clinical-Interview for DSM-IV-TR (SCID-IV) (Wittchen, Wunderlich, Gruschwitz, & Zaudig, 1997). Based on the SCID-IV diagnosis at T_1_ and T_2_ participants were categorized as HC, MDD congruently remitted (i.e. not fulfilling diagnostic criteria for current MDD) or congruently acute at T_1_ and T_2_ (MDD_C_), or changing from acute to remitted or vice versa from T_1_ to T_2_ (incongruent, MDD_IC_).

### Statistical analyses

Analysis scripts and data sets are publicly available (https://osf.io/9754h/) as this study was a secondary preregistration (for details see the supplementary methods). An a priori significance threshold of *p*<.05 was defined and all analyses were conducted in SPSS (IBM) version 26.

Two-year test-retest reliability of the continuous CTQ scores was assessed using absolute agreement in random two-way mixed effect intraclass correlation coefficients (ICC) (Qin, Nelson, McLeod, Eremenco, & Coons, 2019). Cohen’s Kappa (Cohen, 1960) was used for the reliability analysis of categorical scoring (maltreated vs. non-maltreated). According to our first hypothesis, the test-retest reliability was expected to be lower in groups with MDD symptoms as compared to the HC group, and lowest within the group of MDD patients that had incongruent remission states at T_1_ and T_2_ (MDD_IC_). For all MDD patients ΔCTQ was predicted in regression models by ΔBDI, BDI_T1_, age_T1_, and the ΔBDI*BDI_T1_-interaction using a stepwise approach. This analysis allows an estimation of the extent to which a longitudinal change in continuous CTQ scores is systematically associated with changes in longitudinal BDI scores, testing the hypothesis that retrospective self-reports of CM are biased in the direction of higher scores associated with greater depression severity. Further, the regression allows the investigation whether T_1_ depression severity or age of the participants can be used to predict the CTQ stability. The stepwise regression approach eliminates non-informative variables, while informative variables can be identified.

Analyses were initially conducted in the MACS discovery sample in order to derive a statistical model for an adjustment procedure of CM reports. Subsequently, we used the regression weights of the analysis in our discovery sample to correct CM scores in our validation sample (MNC). We then investigated the benefits for the test-retest reliability of the CM assessments of the validation sample when applying the prior correction procedure (second hypothesis).

To assess the robustness of findings, analyses were repeated excluding participants with scores in minimization denial >0, and excluding overly influential cases for the regression models (a priori defined as cases with a Cook’s distance exceeding three standard deviations).

Further, analyses were repeated with the five CTQ subscale scores as dependent variables, as well as with HDRS ratings in order to identify differences in temporal stability across maltreatment subtypes, and to investigate if self-reported depression symptoms affect CTQ reports differently as compared to clinician rated symptom severity. To investigate the susceptibility of categorical CTQ scoring for depressive negativity bias, analyses were repeated using multinomial logistic regressions.

## Results

### Temporal stability of maltreatment reports

Descriptively, individual reports of CM varied on average 3.77 (SD 3.91) and 4.02 (SD 4.19) CTQ sum points between T_1_ and T_2_ in the two cohorts (absolute change). Changes in scores during the interval ranged up to 22 CTQ points. Variability metrics are presented in Table S1.

Analysis of the CTQ sum scores yielded an excellent two-year test-retest reliability with ICC=.956 for the overall MACS sample. Subgroup analyses yielded highly similar reliability estimates for HC, MDD_C_, and MDD_IC_ samples (Table 1, Fig 1A and 1C). The five subscales of the CTQ also showed high reliabilities overall. While its reliability was still high (ICC=.836), the physical neglect subscale had a significantly lower reliability compared to other subscales (Table S2).

**Table 1.** Test-retest reliability estimates of continuous and categorical childhood maltreatment across MACS and MNC subsamples.

|  | Continuous maltreatment |  |  |  | Categorical maltreatment |  |  |  |
| --- | --- | --- | --- | --- | --- | --- | --- | --- |
| | ICC | 95% CI | | *p* | Cohen's $\kappa$ | 95% CI | | *p* |
|  |  | lower | upper |  |  | lower | upper |  |
| MACS |  |  |  |  |  |  |  |  |
| Full sample | .956 | .949 | .963 | <.001 | .551 | .479 | .622 | <.001 |
| HC | .919 | .900 | .935 | <.001 | .383 | .272 | .488 | <.001 |
| MDD<sub>C</sub> | .953 | .938 | .965 | <.001 | .549 | .419 | .671 | <.001 |
| MDD<sub>IC</sub> | .946 | .917 | .965 | <.001 | .554 | .348 | .725 | <.001 |
| MNC |  |  |  |  |  |  |  |  |
| Full sample | .950 | .933 | .963 | <.001 | .507 | .371 | .640 | <.001 |
| HC | .913 | .868 | .943 | <.001 | .360 | .139 | .568 | <.001 |
| MDD<sub>C</sub> | .908 | .804 | .957 | <.001 | .307 | -.149 | .703 | .096 |
| MDD<sub>IC</sub> | .970 | .947 | .984 | <.001 | .568 | .311 | .786 | <.001 |

*Note.* 95% confidence intervals (CI) for reliability estimates of categorical maltreatment are calculated using bootstrapping (N=1000 samples). HC, healthy controls; MDD, major depressive disorder participants with congruent (MDD<sub>C</sub>) or incongruent (MDD<sub>IC</sub>) remission status across T<sub>1</sub> and T<sub>2</sub>.

**Fig 1.**
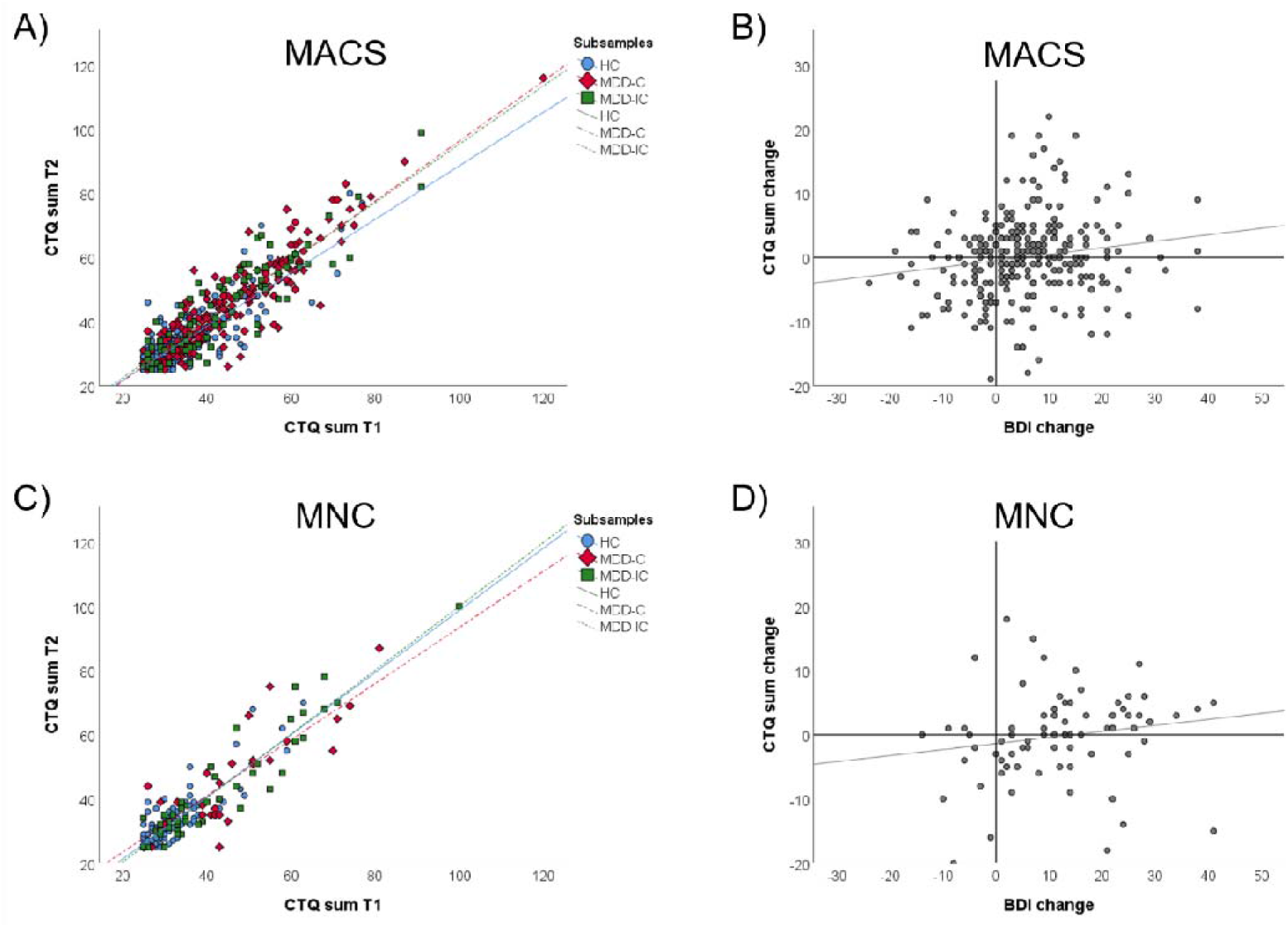
Association of CTQ sum scores at T_1_ and T_2_ (panel A and C) and relationship between CTQ and BDI changes over a 2-year interval among MDD patients (panel B and D) across MACS and MNC samples.

Conducting test-retest analysis with categorical classification of CM yielded moderate agreement between T_1_ and T_2_ (K=.551). Again, reliabilities did not differ between HC, MDD_C_, and MDD_IC_. Analyses yielded highly consistent reliability estimates across cohorts and variables (i.e. continuous, categorical, and CTQ subscale analyses; Table 1 and Table S2).

### Predictors of temporal stability of maltreatment reports

In the MDD subsample of the MACS, ΔBDI significantly predicted ΔCTQ (n=271, *b*=0.101, 95%-CI[.025, .177], *p*=.009, R^2^=.021) indicating that an increase in depression severity over time was associated with an increase in reports of CM. The predictors BDI_T1_, age, and the interaction between ΔBDI and BDI_T1_ were excluded from the model due to insufficient predictive value (all *p*>.165). This predictive effect of ΔBDI could not be replicated in the MNC sample (ΔBDI: *b*=0.292, *p*=.320; all other effects: *b*<|0.130|, *p*>.446). The association between ΔCTQ and ΔBDI within all MDD participants across both cohorts is presented in Fig 1B and 1D. Changes in depression were mainly related to changes in emotional maltreatment subscales (Table S3).

Conducting the logistic regression for categorical maltreatment classification (maltreated vs. non-maltreated), or using the HDRS instead of the BDI, consistently yielded non-significant predictive effects of all variables in both cohorts. Further, results did not change qualitatively in several sensitivity analyses when excluding influential cases or cases with high minimization/denial scores (see supplementary results).

In order to follow up on the weak but significant association between ΔBDI and ΔCTQ in the MACS cohort and the trend in the MNC cohort, exploratory investigations of CTQ subscales were conducted. These analyses revealed that ΔBDI was associated with changes in emotional neglect in the MACS cohort and with emotional abuse in the MNC cohort but not with changes in other CTQ subscales (see supplementary results).

Adjusting CTQ scores in the MNC validation sample for weighted BDI scores based on regression weights obtained in the MACS sample did not yield a change in test-retest reliability of CTQ scores (see supplementary results).

### Maltreatment reports of healthy controls with initial depressive episode in interval

A sample of diagnosis converters was investigated in an additional exploratory analysis. These converters were defined as participants with no history of psychiatric condition at T_1_ but who fulfilled the criteria for an MDD diagnosis during the interval to T_2_. Within MACS and MNC combined, a total of n=27 of such converters was available that were jointly analyzed. The analysis yielded a high temporal stability of CTQ sum scores (ICC=.933, 95%-CI[.853, .969], *p*<.001) with overlapping confidence intervals as for HC participants from both cohorts that did not experience depression symptoms during the interval (Fig 2). Similarly, ΔCTQ was not associated with ΔBDI (*b*=0.021, *p*=.933) in this sample in the regression model.

**Fig 2.**
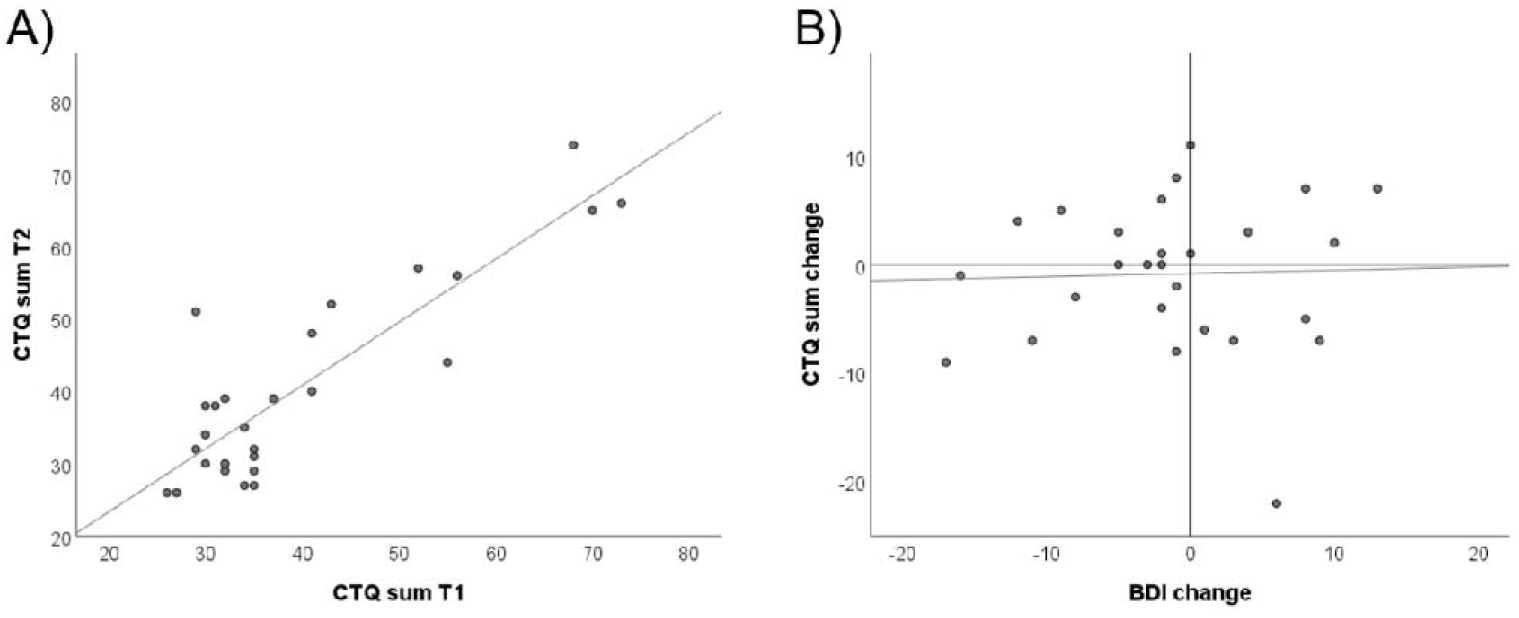
Panel displays A) association of CTQ sum scores at T_1_ and T_2_, and B) association of changes in the CTQ with changes in the BDI in the converter sample.

### Cross-sectional associations between maltreatment reports and depression severity

To follow up on planned longitudinal analyses, we conducted exploratory analysis to examine the relationship between baseline CTQ scores (at T_1_) with BDI scores of the MDD sample at both time points. Spearman correlations indicated that CTQ_T1_ was significantly associated with BDI_T1_ (*r*=.197, 95%-CI[.075, .312], *p*=.001) and BDI_T2_ (*r*=.298, 95%-CI[.183, .401], *p*<.001) in MDD participants of the MACS sample. Within the MNC MDD sample CTQ_T1_ was also significantly associated with BDI_T1_ (*r*=.318, 95%-CI[.092, .522], *p*=.005), however, the association with BDI_T2_ failed to reach significance (*r*=.170, 95%-CI[-.057, .385], *p*=143).

## Discussion

The primary findings of this investigation are that adult retrospective reports of CM using the CTQ are highly temporally stable over a two-year interval and that they are only marginally susceptible to selective negative bias in depressed patients. Temporal stability of continuous maltreatment load was superior to categorical classification of maltreatment.

Retrospective self-reports of CM have been frequently questioned due to potential memory or reporting biases particularly due to depressive symptomatology (Hardt & Rutter, 2004). Reports of low overlap between retrospective and prospective maltreatment measures have further nourished the notion of unclear validity of retrospective assessment instruments (Baldwin et al., 2019). Thus, we aimed to estimate the extent to which such biases occur in a longitudinal design. We found that temporal stability of retrospective maltreatment reports is largely identical between healthy controls and patients with depression. Furthermore, test-retest reliability of CTQ scores was equally high in MDD patients with shifting remission states (from acute to remitted or vice versa) as compared to congruent remission states at both assessment time points. These findings suggest that the temporal stability of retrospective maltreatment reports is not lower per se in people with a lifetime depressive disorder or with acute depressive symptomatology. This notion is largely consistent with a previous study that found equally stable maltreatment reports between clinical and non-clinical samples, using a non-validated interview (Fergusson et al., 2000). Although not directly probing maltreatment experiences, an early study investigating parental bonding recall, similarly found that individuals’ reporting behavior was highly comparable whether depression symptoms were acute or remitted (Parker, 1981).

Investigating intra-individual changes in depressive symptoms we did observe that longitudinal shifts towards higher depression severity were associated with higher levels of reported maltreatment experiences in our larger sample (n=271). However, this effect was small, with changes in depressive symptoms accounting for only 2% of variance in shifts of maltreatment reports. Also, this association failed to reach significance in an independent smaller sample, presumably due to lower statistical power given the small effect size.

Despite little evidence for a depressive recall bias in the above analysis, one could still argue that participants suffering from a depression at both time points reliably show stably altered reporting of maltreatment experiences, without causing increased variability in reports. The notion of a lifetime depression causing a temporally stable bias is consistent with findings of cognitive biases, as overgeneralization of one’s own past, persisting even after remission (Mackinger, Pachinger, Leibetseder, & Fartacek, 2000). A particular strength of the current study is the inclusion of a sample with participants that had an initial depressive episode between both measurement time points (n=27). Data from this sample suggest that adults that have never experienced clinically relevant depressive symptoms do not start reporting on their childhood experiences differently once experiencing an initial depressive episode. While this finding further strengthens the validity of the CTQ measure, low statistical power for this particular sample limits conclusions about a potential small-scale effect of depressive symptoms on maltreatment reports in adults with an initial depressive episode. Mixed findings regarding a depressive bias on retrospective maltreatment reports have been reported by previous studies. Evidence from a large epidemiological study indicates that 12-year test-retest incongruence in reports of adverse experiences can be predicted by depression, and psychological distress (Colman et al., 2016). However, a non-validated interview about a wide variety of experiences (e.g. hospital stay, parental unemployment, physical abuse) was used in this study making it difficult to generalize results to other maltreatment measures. In another epidemiological study also using a non-validated interview, the authors concluded that in a three-year interval a compound mental health index (implicating various symptom domains) accounted for a minor amount of the variability of sexual abuse reports but not for variability in physical abuse assessments (Fergusson, Horwood, & Boden, 2011). No evidence for a recall bias in any abuse or neglect subscales was found by Pinto and colleagues who investigated stability of maltreatment reports via a questionnaire similar to the CTQ in a six-month interval in institutionalized young adults (Pinto, Correia, & Maia, 2014). Differences in assessments methods, as well as sample characteristics of these studies make a comparison difficult. However, taken together with our findings the evidence suggests that a reporting bias due to depressive symptomatology is not present or negligible due to very small effect sizes. This seems to be the case particular for retrospective maltreatment reports assessed via a well-validated measure as the CTQ. While the stability of the total score of the CTQ was largely independent of depressive symptom change, our findings suggest that reports of emotional types of maltreatment may tend to be more susceptible to depressive symptoms as compared to other subtypes of abuse and neglect. Thus, clinicians and researchers utilizing emotional subscales of the CTQ as isolated measures are advised to interpret the reports with caution regarding a potential bias.

We originally aimed to test an adjustment procedure that corrects CTQ scores for current depressive symptom severity in order to improve the validity of the retrospective maltreatment reports. However, this approach did not yield a psychometric improvement which seems reasonable considering that findings suggest low susceptibility of CTQ scores for depressive symptoms, and the temporal stability was already excellent prior to any correction procedure. Baseline depression severity and age were also not predictive for variability in CTQ scores.

Overall, we found CTQ scores to be highly temporally stable across a two-year interval. Previous studies investigating the temporal stability of various retrospective maltreatment measures other than the CTQ mostly reported much lower test-retest estimates particularly when a categorization of maltreatment experiences was implemented. Congruence no better than chance between interview reports of autobiographical childhood memories (including physical and emotional punishment) was reported by Offer et al. (2000). Moderate congruence of retrospective reports on sexual and physical abuse were presented in a community sample (Fergusson et al., 2000), and a mixed psychiatric sample (Hardt et al., 2006). In contrast test-retest agreement of continuous abuse and neglect scores based on questionnaire data of institutionalized adolescents and young adults was found to be more comparable to our results although somewhat lower (Pinto et al., 2014). When only regarding research investigating the CTQ, the reported temporal stability of continuous (Bernstein et al., 1994; Shannon et al., 2016) and categorical (Cammack et al., 2017; Shannon et al., 2016) data is highly similar to our estimates. Notably, evidence consistently indicates that temporal stability of continuous CTQ scores tends to be higher as compared to the stability of categorical classifications. Superiority of continuous over categorical assessments is well-documented concerning various psychopathological measures (Markon, Chmielewski, & Miller, 2011).

We further replicated previous findings that CM experiences are associated with more severe depressive symptoms (Lippard & Nemeroff, 2020). Baseline CTQ scores associated with concurrent depression severity and predictive for symptomatology two years later. Considering that we found little evidence for a depressive bias on the reports of CM this supports the notion of CM experiences negatively affecting psychopathology later in life.

There are several important limitations. The generalization to other measures of retrospective self-reports of maltreatment is unclear. Further, our study design is not sensitive for potential long-term effects of chronic depressive symptoms over the course of more than two years on the reporting of CM. Notably, while on average CTQ scores were temporally stable, some individuals showed very high fluctuation in maltreatment reports. A potential reporting bias may be moderated by other factors that were not regarded in the current study as specific comorbidities or personality profiles. Further, so-called ‘biases’ e.g. due to depression may be brought about by increased administration of psychotherapy that may cause a more accurate reflection on one’s own childhood and thereby change appraisal and reports of maltreatment.

## Conclusion

In summary, findings from the presented preregistered analyses suggest that the CTQ is a temporally stable tool for the retrospective assessment of CM that is not susceptible for changes in depressive symptoms. The evidence against depression bias in combination with high practicability encourages the utilization of validated retrospective self-report tools for the assessment of CM for researchers and clinicians. Further, findings promote a continuous rather than categorical conceptualization of maltreatment experiences. Such gradual approach seems to improve psychometric qualities and may also more accurately represent the complex neurobiological basis, as well as clinical consequences of abuse and neglect. Personalized medicine approaches modelling disease trajectories based on retrospectively collected risk variables are therefore highly advised to conceptualize a gradual maltreatment load in order to increase its predictive potential.

## Supporting information

Supplemental Methods and Results

## Data Availability

The data that support the findings of this study are openly available in the Open Science Framework at https://osf.io/9754h/, DOI: 10.17605/OSF.IO/9754H.

## Key points and relevance

### What’s known?

- In cross-sectional studies, depression and childhood maltreatment (CM) experiences are often associated, which is why it was suggested that retrospective reports of CM could be biased by depressive symptoms

### What’s new?

- Temporal stability of CM reports is independent from a diagnosis of depression or its remission state.
- Initially healthy adults do not report on their childhood experiences differently once experiencing a first depressive episode.
- Reports of emotional types of maltreatment may be more susceptible to depressive bias as compared to other subtypes of abuse and neglect.

### What’s relevant?

- The CTQ can be used to assess retrospective reports of CM in clinical cohorts with high temporal stability, making it a useful screening tool in clinical practice.

## Acknowledgments

This work is part of the German multicenter consortium “Neurobiology of Affective Disorders. A translational perspective on brain structure and function”, funded by the German Research Foundation (Deutsche Forschungsgemeinschaft, DFG; Forschungsgruppe/Research Unit FOR2107). DFG funding in the context of the FOR2107 was received by: TK (speaker FOR2107; DFG grant numbers KI 588/14-1, KI 588/14-2, KI 588/15-1, KI 588/17-1), UD (co-speaker FOR2107; DA 1151/5-1, DA 1151/5-2, DA 1151/6-1), AK (KR 3822/5-1, KR 3822/7-2), IN (NE 2254/1-2), TH (HA 7070/2-2, HA7070/3, HA7070/4). This work was further supported by the Interdisciplinary Center for Clinical Research (IZKF) of the medical faculty of Münster (Grant Dan3/012/17 to UD and SEED 11/19 to NO). EL was additionally supported by the Christiane Nüsslein-Vollhard Foundation.

## Data access and responsibility

All PIs take responsibility for the integrity of the respective study data and their components. All authors and co-authors had full access to all study data.

## Abbreviations

CTQ: Childhood Trauma Questionnaire
HC: healthy controls
MACS: Marburg-Münster Affective Disorders Cohort Study
MDD: major depressive disorder
MNC: Münster Neuroimaging Cohort

