## Supplemental Methods and Results for "Temporal stability and state-dependence of retrospective self-reports of childhood maltreatment in major depression: a two-year longitudinal analysis of the childhood trauma questionnaire"

**Online Supplements**

*Supplementary methods*

*Supplementary results*

*Supplementary tables and figures*

**Table S1.** Descriptive statistics for the individual temporal variation of CTQ and BDI scores over a two-year interval.

**Table S2.** Test-retest reliability of continuous maltreatment load in CTQ subscales across MACS and MNC subsamples.

**Table S3.** Correlations between BDI change and change in CTQ subscales in the MACS and MNC samples.

**Figure S1.** Flowchart of the data exclusion steps for the MACS and MNC samples.

*Supplementary methods*

As both cohorts were taken from ongoing longitudinal neuroimaging and genotyping studies, additional exclusion criteria for both cohorts and both time points were related to MRI and genetic assessments. Exclusion criteria included general MRI contradictions, color blindness, severe physical impairment (e.g. cancer, epilepsy), head trauma or unconsciousness, history of seizures, neurological conditions, non-caucasian origin or general non-compliance with the study protocol. Participants that converted from an MDD diagnosis to a bipolar diagnosis from T_1_ to T_2_ were excluded. We further excluded participants with a lifetime diagnosis of schizophrenia, schizoaffective disorder, bipolar disorder, or substance dependence from both samples. Only participants with complete assessment at T_1_ and T_2_ were included.

Hypotheses and analysis steps were planned and preregistered a priori before the sighting of the data, as a secondary preregistration (van den Akker et al., 2019). Data collection and preprocessing took place before the preregistration by researchers who were blind to the current hypotheses. Analysis scripts and data sets are publicly available (<https://osf.io/9754h/)>. Deviations from preregistered protocols are fully disclosed within the same repository and non-planned analyses designated as exploratory. Preregistration addresses problems of replicability and likely minimizes publication and researcher bias by reducing researcher degrees of freedom (Allen & Mehler, 2019).

*Supplementary results*

Using HDRS ratings instead of BDI scores the stepwise regression yielded a non-significant effect of ΔHDRS on ΔCTQ in the MACS sample (n=271, *b*=0.188, *p*=.117), and the MNC sample (n=76, *b*=.274, *p*=.512).

For logistic regression analyses predicting congruence of categorical maltreatment classification, three outcome groups were defined: 1) congruent classification of maltreatment over T_1_ and T_2_ (C), 2) incongruent classification with maltreated at T_1_ and non-maltreated at T_2_ (IC_lower_), and 3) incongruent classification with non-maltreated at T_1_ and maltreated at T_2_ (IC_higher_). Multinomial logistic regression in the MDD sample within MACS also yielded non-significant effects of ΔBDI (χ²(2)=1.495, *p*=.473), BDI_T1_(χ²(2)=1.308, *p*=.520), age (χ²(2)=0.853, *p*=.653), and the interaction between ΔBDI and BDI_T1_ (χ²(2)=0.562, *p*=.755). Comparable results were obtained in the MNC sample with all non-significant predictors in an identical logistic regression model (all *p*>.349).

Regression analyses yielded qualitatively identical results when excluding participants with minimization/denial scores exceeding zero at T1 or T2 (MACS: n=51 MDD cases, MNC: n=28 MDD cases), and when excluding participants with a Cook’s distance of >3SD over the mean (MACS: >0.021; n=10 MDD cases; MNC: >0.159; n=1).

When correcting the CTQ score in the MNC sample with the weighted BDI based on results obtained in the MACS sample (CTQ_T1corr_ = CTQ_T1_ - 0.101* BDI_T1_, CTQ_T2corr_=CTQ_T2_ - 0.101*BDI_T2_) the reliability slightly decreased for the MDD subsample (n=76, ICC=.947, 95%-CI [.916, .967], *p* <.001). However, as the confidence intervals of reliabilities prior and after correction overlap, this decrease was not significant, indicating that the adjustment of CTQ scores for concurrent depression severity did not yield an improvement of its temporal stability.

**Table S1.** Descriptive statistics for the individual temporal variation of CTQ and BDI scores over a two-year interval.

|  | CTQ change | | | |  | BDI change | | | |
| --- | --- | --- | --- | --- | --- | --- | --- | --- | --- |
|  | mean | SD | median | range |  | mean | SD | median | range |
| MACS |  |  |  |  |  |  |  |  |  |
| Full sample (n=598) | 0.174 | 5.429 | 0 | [-20; 22] |  | 3.005 | 7.535 | 1 | [-24; 38] |
| HC (n=327) | 0.269 | 4.575 | 0 | [-20; 19] |  | 1.000 | 3.840 | 1 | [-13; 24] |
| MDD-C (n=188) | 0.218 | 6.121 | 0 | [-19; 22] |  | 3.447 | 8.713 | 3 | [-19; 38] |
| MDD-IC (n=83) | -0.301 | 6.757 | 0 | [-14; 16] |  | 9.916 | 10.804 | 10 | [-24; 38] |
| MNC |  |  |  |  |  |  |  |  |  |
| Full sample (n=168) | -0.548 | 5.804 | 0 | [-20; 18] |  | 4.982 | 10.748 | 0 | [-14; 41] |
| HC (n=92) | -0.815 | 4.638 | 0 | [-17; 12] |  | -0.935 | 2.843 | 0 | [-10; 41] |
| MDD_C_ (n=29) | -0.241 | 8.919 | 0 | [-20; 18] |  | 8.138 | 14.235 | 5 | [-14; 41] |
| MDD_IC_ (n=47) | -0.213 | 5.568 | 0 | [-15; 12] |  | 14.617 | 10.437 | 14 | [-9; 8] |

*Note.* Positive change scores indicate greater values at T_1_ as compared to T_2_ and negative scores vice versa. CTQ, childhood trauma questionnaire; BDI, Beck Depression Inventory; MACS, Marburg-Münster Affective Disorders Cohort Study; MNC, Münster Neuroimaging Cohort; HC, healthy controls; MDD, major depressive disorder with congruent (C) or incongruent (IC) remission status.

**Table S2.** Test-retest reliability of continuous maltreatment load in CTQ subscales across MACS and MNC subsamples.

|  | MACS | | | |  | MNC | | | |
| --- | --- | --- | --- | --- | --- | --- | --- | --- | --- |
|  |  | 95% CI | |  |  |  | 95% CI | |  |
|  | ICC | Lower bound | Upper bound | p |  | ICC | lower bound | upper bound | *p* |
| *Emotional abuse* |  |  |  |  |  |  |  |  |  |
| Full sample | .925 | .911 | .936 | <.001 |  | .924 | .897 | .944 | <.001 |
| HC | .871 | .840 | .896 | <.001 |  | .883 | .823 | .923 | <.001 |
| MDD-C | .925 | .900 | .943 | <.001 |  | .887 | .760 | .947 | <.001 |
| MDD-IC | .893 | .835 | .931 | <.001 |  | .932 | .878 | .962 | <.001 |
| *Physical abuse* |  |  |  |  |  |  |  |  |  |
| Full sample | .935 | .924 | .945 | <.001 |  | .935 | .911 | .952 | <.001 |
| HC | .904 | .880 | .923 | <.001 |  | .924 | .885 | .950 | <.001 |
| MDD-C | .936 | .915 | .952 | <.001 |  | .921 | .831 | .963 | <.001 |
| MDD-IC | .952 | .926 | .969 | <.001 |  | .943 | .898 | .968 | <.001 |
| *Sexual abuse* |  |  |  |  |  |  |  |  |  |
| Full sample | .932 | .920 | .942 | <.001 |  | .891 | .852 | .919 | <.001 |
| HC | .915 | .895 | .932 | <.001 |  | .964 | .945 | .976 | <.001 |
| MDD-C | .952 | .937 | .964 | <.001 |  | .815 | .610 | .913 | <.001 |
| MDD-IC | .835 | .744 | .893 | <.001 |  | .902 | .824 | .946 | <.001 |
| *Emotional neglect* |  |  |  |  |  |  |  |  |  |
| Full sample | .932 | .921 | .942 | <.001 |  | .939 | .917 | .955 | <.001 |
| HC | .903 | .879 | .922 | <.001 |  | .898 | .846 | .932 | <.001 |
| MDD-C | .904 | .872 | .928 | <.001 |  | .937 | .866 | .971 | <.001 |
| MDD-IC | .933 | .897 | .957 | <.001 |  | .952 | .913 | .973 | <.001 |
| *Physical neglect* |  |  |  |  |  |  |  |  |  |
| Full sample | .836 | .807 | .860 | <.001 |  | .815 | .750 | .864 | <.001 |
| HC | .618 | .525 | .693 | <.001 |  | .688 | .530 | .794 | <.001 |
| MDD-C | .858 | .811 | .894 | <.001 |  | .813 | .599 | .913 | <.001 |
| MDD-IC | .883 | .816 | .925 | <.001 |  | .894 | .810 | .941 | <.001 |

*Note.* MACS, Marburg-Münster Affective Disorders Cohort Study; MNC, Münster Neuroimaging Cohort; CI, confidence interval; ICC, intraclass correlation; HC, healthy controls; MDD, major depressive disorder with congruent (C) or incongruent (IC) remission status.

**Table S3.** Correlations between BDI change and change in CTQ subscales in the MACS and MNC samples.

|  | ΔBDI | |
| --- | --- | --- |
|  | MACS | MNC |
| ΔEA | .027 | .240^*^ |
| ΔPA | -.023 | -.047 |
| ΔSA | .073 | .051 |
| ΔEN | .221^**^ | .090 |
| ΔPN | .094 | .076 |

*Note.* Pearson correlations based on major depressive disorder subsamples are presented. Change scores are calculated as T_1_-T_2_. BDI, Beck Depression Inventory; EA, emotional abuse; PA, physical abuse; SA, sexual abuse; EN, emotional neglect; PN, physical neglect. *p<.05, **p<.001.

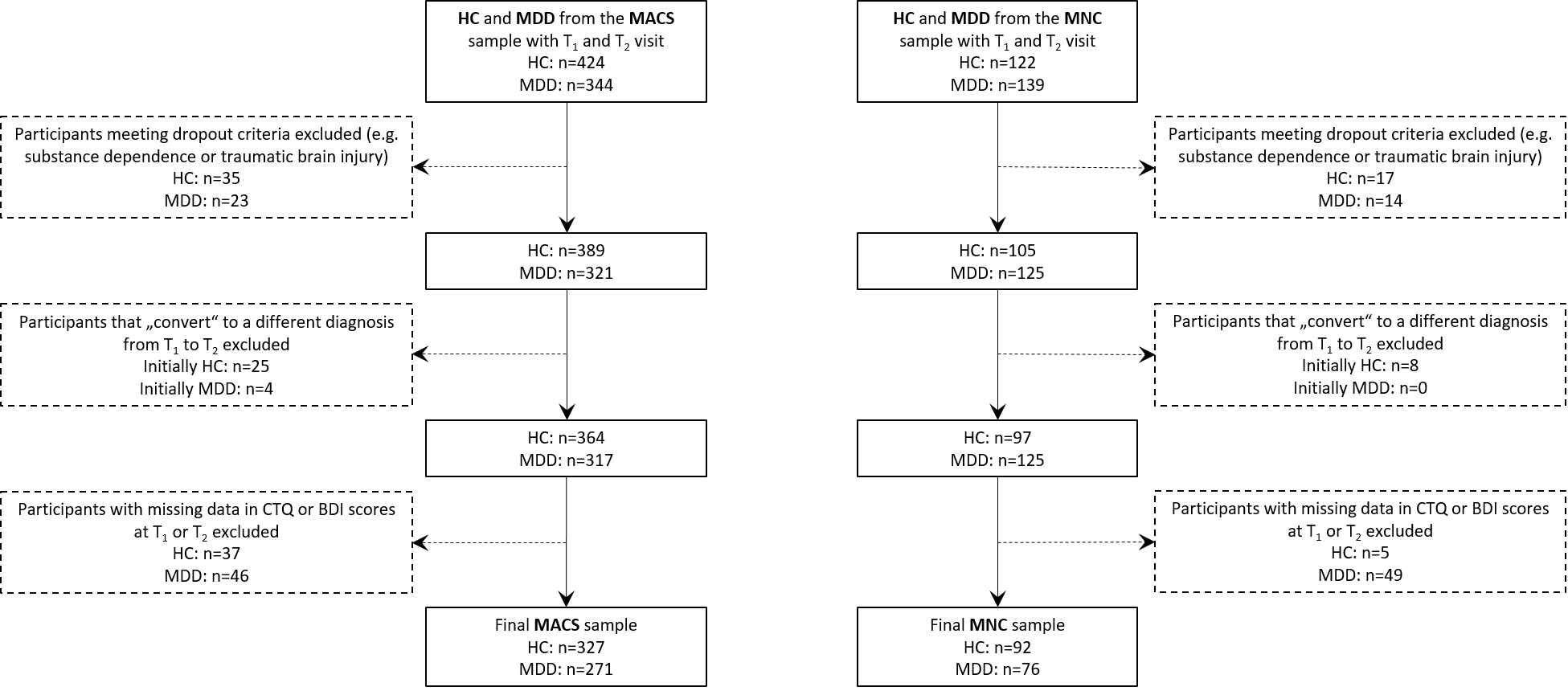

**Figure S1.** Flowchart of the data exclusion steps for the MACS and MNC samples.
